# Long-Term Outcomes of SARS-CoV-2 Variants and Other Respiratory Infections: Evidence from the Virus Watch Prospective Cohort in England

**DOI:** 10.1101/2023.12.18.23300124

**Authors:** Sarah Beale, Alexei Yavlinsky, Wing Lam Erica Fong, Vincent G Nguyen, Jana Kovar, Theo Vos, Sarah Wulf Hansen, Andrew C Hayward, Ibrahim Abubakar, Robert W Aldridge

## Abstract

**Background:** Given the considerable prevalence of long-term sequelae following SARS-CoV-2 infection, understanding pathogen-related factors that influence long-term outcomes is warranted. We aimed to compare the likelihood of long-term symptoms for SARS-CoV-2 variants, other acute respiratory infections (ARIs) and non-infected individuals.

**Method:** Data were from 5,630 individuals participating in Virus Watch, a prospective community cohort study of SARS-CoV-2 epidemiology in England. We used logistic regression to compare the predicted probability of developing long-term symptoms (>2 months duration) during different variant dominance periods according to infection status (SARS-CoV-2, other ARI, or no infection), adjusting for confounding by demographic and clinical factors and vaccination status.

**Results:** Predicted probability of long-term sequelae was greater following SARS-CoV-2 infection during the Wild Type (adjusted predicted probability (PP) 0.28, 95% confidence interval (CI) =0.14-0.43), Alpha (PP= 0.28, 95% CI =0.14-0.42), Delta (PP= 0.34, 95% CI=0.25-0.43) and Omicron BA.1 periods (PP= 0.27, 95% CI =0.22-0.33) compared to later Omicron sub-variants (PP range from 0.11, 95% CI 0.08-0.15 to 0.14, 95% CI 0.10-0.18). While differences between SARS-CoV-2 and other ARIs (PP range 0.08, 95% CI 0.04-0.11 to 0.23, 95% CI 0.18-0.28) varied by period, estimates for long-term symptoms following both infection types substantially exceeded those for non-infected participants (PP range 0.01, 95% CI 0.00,0.02 to 0.03, 95% CI 0.01-0.06) across all variant periods.

**Conclusions:** Between-variant differences influenced the likelihood of post-infection sequelae for SARS-CoV-2, with lower predicted probabilities for recent Omicron sub-variants similar to those for other contemporaneous ARIs. Both SARS-CoV-2 and other ARIs were associated with long-term symptom development, and further aetiological investigation including between-pathogen comparison is recommended.

## Background

The Coronavirus Disease 2019 (COVID-19) pandemic has left an indelible mark on global population health, not only through widespread acute illness and mortality but also through disabling chronic symptoms that affect a substantial portion of those previously infected with Severe Acute Respiratory Syndrome Coronavirus 2 (SARS-CoV-2). The World Health Organization (WHO) defines Post-Covid Condition (PCC), also commonly known as Long Covid, as the persistence or development of new symptoms within three months of SARS-CoV-2 infection that last for at least two months and have no alternative explanation(1,2).

Estimates of PCC incidence following acute infection vary substantially, with 10-30% of people with mild-to-moderate infections and over 50% of those with severe infections estimated to develop persistent long-term symptoms (1,3). Diverse symptoms affecting a range of organ systems have been reported, with common symptoms including fatigue, shortness of breath, palpitations, cognitive dysfunction, and joint and/or muscle pain (4). These symptoms have notable overlap with long-term symptoms reported after other respiratory viral infections and with chronic conditions such as myalgic encephalomyelitis/chronic fatigue symptom (ME/CFS), which may have post-infectious onset (5,6).

While the aetiology of PCC is under investigation, evidence supports several potential mechanisms including viral persistence and reactivation of latent viruses, pathologies of the inflammatory response and autoimmunity, and coagulopathies and endothelial dysfunction, which may interact and produce different symptoms depending on the organ system(s) affected (1,7). Epidemiological investigation into factors associated with risk of PCC is relevant both to inform and respond to growing aetiological understanding of the condition and to provide evidence around vulnerable groups to inform public health responses. Risk-relevant factors are likely to reflect features of both the pathogen and the infected person, and established factors include female sex, obesity, and long-term conditions affecting the immune system (7,8). SARS-CoV-2 variant may be an important pathogen-related determinant of PCC risk, given different viral loads and systemic impacts associated with different variants (5); however, understanding of its impact on the development of PCC is relatively limited.

Several studies representing a range of global regions have investigated the likelihood of developing long-term sequelae following COVID-19 infection according to variant of infection, with studies consistently finding a lower likelihood of long-term sequelae following infections with the Omicron variant compared to the ancestral Wild Type strain or subsequent variants of concern (VoCs) (9–20). There is some evidence that Wild Type infections were associated with a greater likelihood of developing long-term sequelae compared to subsequent variants (9,12,16,21), though between-variant differences prior to Omicron are less consistent across studies. However, current understanding of the impact of SARS-CoV-2 variant on likelihood of post-acute sequelae are impacted by several methodological concerns. The definition of post-acute sequelae is highly variable across studies, and often relies on a relatively short duration of follow-up (e.g., symptoms persisting within a month of infection). Echoing a common issue within PCC research more broadly, many study samples are based on hospitalised patients, which is likely to impact between-variant differences given greater overall severity associated with hospitalisation and also limit generalisability and consequent ability to inform public health decision making. Furthermore, many current estimates have limited or no adjustment for potential confounding (9), or rely on stepwise selection procedures or mutually-adjusted models designed to evaluate a range of exposures. Unadjusted or under-adjusted estimates are likely to be substantially impacted by confounding by demographic and clinical factors or vaccination status, while estimates that are not adjusted with specific consideration for variant as the exposure may be impacted by incorrect specification. Finally, mutations in the Omicron variant have given rise to a range of sub-variants which are regarded as independent VoCs and have become responsible for all current SARS-CoV-2 infections (22); however, delineated comparisons including Omicron sub-lineages are currently lacking.

Long-term symptoms following viral infections are not unique to SARS-CoV-2, and have been identified after other common acute respiratory infections (ARIs) such as influenza, rhinovirus, and respiratory syncytial virus (6,23). Comparison of post-acute sequelae following SARS-CoV-2 versus other ARIs is very limited, particularly in general population samples. One primarily USA-based study of electronic health records found evidence of post-acute sequelae following both COVID-19 and influenza illness in 2020, with some evidence of a higher recorded likelihood following COVID-19 (24). A further UK-based study found that people who experienced COVID-19 or other ARIs between January-February 2021 had a greater likelihood of experiencing symptoms >12 weeks post-infection compared to contemporaneous non-infection controls, with the probability of some symptoms such as disruption to smell and taste was greater following COVID-19 than other ARIs (25). Comparisons between a range of SARS-CoV-2 variants and other ARIs are currently unavailable, and are warranted given evidence of differential likelihood of long-term symptoms particularly for more recent variants. Furthermore, contemporaneous comparison with participants who did not experience an infection would provide a valuable benchmark for symptom development given substantial changes across time in COVID-related restrictions, population contact patterns, and other socio-behavioural influences on health.

This analysis aimed to address these gaps in the literature around long-term sequelae of respiratory infections during the COVID-19 pandemic using data from the Virus Watch prospective cohort study in England. Objectives were to:

1. investigate how the risk of PCC varied according to SARS-CoV-2 variant of infection
2. compare how the risk of new-onset long-term symptoms differed between SARS-CoV-2 variants, other acute respiratory infections, and individuals with no detected infection during each variant dominance period.

## Methods

### Ethics Approval and Consent

Virus Watch was approved by the Hampstead NHS Health Research Authority Ethics Committee: 20/HRA/2320, and conformed to the ethical standards set out in the Declaration of Helsinki. All participants provided informed consent for all aspects of the study.

### Participants

Participants (*n*=5,630) were a subset of the Virus Watch longitudinal cohort study (*n* = 58,628). Virus Watch is a community prospective cohort study established in June 2020 and investigates acute infection syndromes and SARS-CoV-2 infections in households across England and Wales. The study involves weekly questionnaires about symptoms, SARS-CoV-2 testing, and vaccinations, as well as bespoke monthly questionnaires investigating demographic, clinical and psychosocial topics relevant to COVID-19 that are beyond the scope of the weekly survey. These monthly surveys include a repeated survey related to new-onset long-term symptoms.

Households were recruited into the Virus Watch study using SMS and postal recruitment supported by general practices and using social media campaigns. Eligibility criteria were residence in England or Wales, household size up to six people (due to survey infrastructure limitations), internet and email access, ability to complete surveys in English, and consent or assent from all household members. Further detail of recruitment and methodology are provided in the study protocol (26) and cohort profile (27) and recruitment dates are illustrated in Supplementary Figure S1. Participants in the current long-term symptoms study comprised Virus Watch participants who met the following further inclusion criteria:

1. resident in England, due to data availability around variant periods,
2. responded to monthly survey(s) related to new-onset long-term symptoms covering the period between February 2020 and March 2023,
3. able to be assigned infection status during variant dominance periods based on SARS-CoV-2 clinical testing records from linkage and study data.

### Exposure

The exposure of interest was SARS-CoV-2 variant dominance period, with the exposure stratified by infection status (SARS-CoV-2 infection, other acute respiratory infection (ARI), or no infection; see Statistical Analyses for further description). Variant dominance periods were derived based on date and national region using previously established methodology (28), with dominance periods defined by the date limits between which over 75% of SARS-CoV-2 infections within each of England’s nine national regions were attributed to each variant based on UK Health Security Agency national surveillance (29,30). The following dominance periods were included during the study period: Wild Type, Alpha, Delta, Omicron BA.1, Omicron BA.2, Omicron BA.5, and Omicron Other, i.e. time during the overall Omicron dominance period where multiple sub-lineages circulated and none were dominant. Please see Supplementary Table S1 for variant period dates.

Infection status during each period was classified using linkage to SARS-CoV-2 test results from UK national records, study-based testing, and self-reported information about testing and acute illness episodes provided in the weekly survey, with detail for each infection status provided below.

#### SARS-CoV-2 Infections

SARS-CoV-2 infections were established based on participants’ first evidence of infection, using the following sources:

1. Polymerase-chain reaction (PCR) or lateral flow (LFT) testing from national linkage or study records, which are indicative of current infection at the time of the test. All participants had results available from linkage and self-report across the study period. Testing was also provided by the Virus Watch study during several periods, with the protocol varying over time (please see ‘Virus Watch Testing and Study Outcomes’ in the Supplementary Materials).
2. Serological testing for anti-nucleocapsid antibody or anti-spike antibody (prior to vaccination), which are indicative of prior infection. Serological testing was used to identify SARS-CoV-2 infections if a date of infection could be estimated for the result due to seroconversion during routine monthly testing. Serological tests were also used to exclude participants with a prior infection where a date could not be estimated (e.g., seropositive upon first test). Please see ‘Virus Watch Testing and Study Outcomes’ for further information about serological testing.

Positive tests recorded in Virus Watch and UK national records during the same 14-day period were matched based on a sliding date window, and the nationally-recorded infection date was used in these cases. Participants whose first evidence of infection was based on serological testing in the absence of a prior negative serological test were excluded, as the variant period of their first infection could not be derived and consequently infection status could not be appropriately allocated for each variant period. Positive SARS-CoV-2 cases were limited to first infections as the majority of cases within Virus Watch comprised first infections, due to limitations with detecting reinfections from serology, and due to the different immune profile of reinfections meriting further, adequately-powered research.

#### Other ARI

Other acute respiratory infection during each variant period comprised participants who reported episode(s) of acute illness comprising respiratory symptom(s) (i.e., fever, cough, chills, runny nose, blocked nose, sneezing, sore throat, shortness of breath, wheezing), tested negative for SARS-CoV-2 during illness, and had no evidence of SARS-CoV-2 infection from any source during the given variant period or within three months of the acute respiratory infection episode (i.e., the period during which any post-acute sequelae may be attributed to a given illness following the WHO consensus definition). Participants who reported loss of taste and/or smell during an infection episode were excluded regardless of SARS-CoV-2 test results to prevent misclassification.

While SARS-CoV-2 results were available from the beginning of the pandemic due to linkage and serological testing, Other ARI was based on participants’ reporting of symptoms as well as test results and consequently were only included from the point where participants joined the study (beginning June 2020).

#### No Infection

Participants were considered to have had no infection during a given variant period if they had no record of a SARS-CoV-2 infection (primary or repeat infection) during each variant period from any clinical source, and had not reported any acute symptom(s) consistent with respiratory (fever, cough, chills, runny nose, blocked nose, sneezing, sore throat, shortness of breath, wheezing, loss or change to smell and/or taste), gastrointestinal (diarrhoea and/or vomiting), or glandular (swollen tonsils and/or cervical lymph nodes) infections during the given variant period.

Classification into this exposure group relied on symptom reporting as well as testing records and consequently was limited to the period after which participants were recruited to the study.

### Outcome

The outcome was binary development of new-onset long term-symptoms (yes/no long-term symptoms), with the data items used to derive the binary outcome described below. As SARS-CoV-2 was the primary focus of this analysis, the definition of long-term symptoms was derived from the World Health Organisation consensus definition for PCC: onset of symptoms lasting at least two months within three months of acute infection, which cannot be explained by another diagnosis (31). Participants were included in this group if at least one symptom was present for at least two months, regardless of the duration of other symptoms. Participants were not included in this analysis if all of their symptoms had occurred for less than two months. Where there was no acute infection present (i.e., the ‘no infection’ exposure group), the requirement for symptoms developing within three months of an infection was removed but other criteria remained the same. Participants were classified as not having long-term sequelae if they completed all surveys - to prevent non-detection of unreported symptoms and consequently misclassification - and never reported new-onset long-term symptoms at any point during the study period.

Data were drawn from a survey regarding new-onset long-term symptoms sent to the Virus Watch cohort. The survey requested participants to indicate whether they experienced any “new symptoms … for four or more weeks [during the survey period] …that are not explained by something else (e.g., pre-existing chronic illness or pregnancy).” Participants then provided further information including selecting all symptoms they experienced from a list (see Supplementary Materials) and providing the onset dates of disruptive symptoms.

Participants indicated whether the symptoms had resolved or were ongoing, and were classified as having PCC-compliant symptoms if the duration between onset and survey date (if ongoing) or resolution date (if resolved) was >2 months. Participants whose symptoms were ongoing but with a duration <2 months were excluded as it was not possible to determine whether their symptoms would resolve. Using date data, development of symptoms within three months of a SARS-CoV-2 or other acute respiratory infection within each variant period was also evaluated. Where there was no acute infection, attribution to a variant period was based on symptom onset within the variant boundary dates. The outcome for this analysis was development of long-term symptoms informed by the WHO consensus definition of PCC, and investigation into specific symptom profiles by variant is the subject of a further analysis beyond the scope of this paper.

The survey was sent to the full cohort four times as part of the Virus Watch monthly surveys: in February 2021, May 2021, March 2022, and March 2023. Participants were asked to report on any new-onset long-term symptoms that occurred during the previous year, except in the May 2021 survey where they recalled since February 2020 (when Covid-19 was first reported in the UK (32) as this survey was conducted to account for any non-response to the February 2021 survey. Consequently, responses cover the full period from February 2020 to March 2023. While there may be some overlap in the survey periods, symptoms and their onset dates were matched so that episodes of long-term symptoms could be tracked across multiple surveys.

### Covariates

Where indicated (see Statistical Analyses), models were adjusted for the following demographic and clinical covariates collected upon study registration: age (0-11, 12-19, 20-39, 40-64, 65+), sex at birth, deprivation (English Indices of Multiple Deprivation Quintile), binary comorbidity status (presence of any condition on the UK NHS/government list denoting extreme clinical vulnerability or clinical vulnerability (33), ethnicity (White British versus other), and body mass index (BMI; underweight <18.5kg/m2, healthy weight 18.5-24.9 kg/m2, overweight 25-29.9 kg/m2, obesity class 1 30-34.9 kg/m2, obesity class 2 35-39.9 kg/m2, and obesity class 3 > 40 kg/m2 (34)). Occupation was classified into the following categories triangulating occupation, exposure risk, and employment status: higher exposure risk occupation, lower exposure risk occupation, retired, not in employment, and unknown/other status. Further details of occupational categories are provided in the Supplementary Materials, and details of the occupational coding process are provided in related Virus Watch analyses (35).

COVID-19 vaccination status was accounted for using two variables: number of doses (0, 1, 2, 3, and 4+) and time since vaccination (described below). Maximum number of COVID-19 vaccination doses was defined based on linkage or self-report data up to seven days prior to infection to allow for a sufficient vaccine response to potentially influence infection, or - if no infection occurred - the maximum number of doses received by the end of the variant period (i.e., the end of follow-up for each exposure period). Time since vaccination was defined as the duration in days between the most recent dose and the date of infection or - if no infection occurred - the end of the variant period, and was included to account for antibody building in the days following vaccination and waning following the peak of protection. Time since vaccination was entered as 0 for unvaccinated participants. To account for the non-monotonic, U-shaped relationship between time since vaccination and associated protection, time since vaccination was entered as a quadratic term in relevant models.

### Statistical Analysis

We used stratified logistic regressions to investigate how infection status (SARS-CoV-2, other ARI, no infection) during each variant period influenced the odds of developing new-onset long-term symptoms. Variant period was entered as the exposure and models were stratified by infection status to obtain estimates of both the impact of SARS-CoV-2 variant on likelihood of developing PCC (Aim 1) and to facilitate comparison by infection status within each variant dominance period (Aim 2) within comparable models with a consistent adjustment set without requiring an interaction term for vaccination which would impair interpretability of findings related to the primary aims.

We developed a directed acyclic graph (DAG) to determine the minimally sufficient adjustment set required to estimate the impact of infection status during each variant period of risk of post-acute sequelae (Supplementary Figure S2); the DAG was considered applicable to all infection status types given that the factors related to exposure to SARS-CoV-2 are relevant to other respiratory infections (versus no infection). The minimally sufficient adjustment set suggested included age, sex, ethnicity, occupation, deprivation, comorbidities and vaccination status; BMI was also included in the adjustment set in the analyses due to its prominent influence on acute infections and development of long-term symptoms (36,37) and to facilitate constructing unbiased sequentially-adjusted models as described below (see Supplementary Figure S2). Models were presented unadjusted, adjusted for demographic and clinical factors (age, sex, ethnicity, occupation, deprivation, comorbidities, and BMI), and additionally adjusted for COVID-19 vaccination status. SARS-CoV-2 vaccination status modifies the risk of acquiring SARS-CoV-2 (dependent on the number of doses and waning due to time since recent dose) and consequently may influence infection status overall by altering the likelihood that an individual be included the other ARI or no infection groups according to our definitions by influencing SARS-CoV-2 infection risk. While this was addressed by stratification, we also included vaccination status in models for the Other ARI and No Infection groups as a counterfactual test of the effect of vaccination in SARS-CoV-2 and to assess evidence of misclassification of SARS-CoV-2 cases.

While participants could only appear during a single variant period for SARS-CoV-2 infections (see Exposure section), they could appear in multiple variant periods for the Other ARI and no infection groups and consequently cluster robust standard errors were applied for relevant models. Results were expressed as predictive probabilities rather than odds ratios to enable comparison across all variant periods rather than in relation to the reference category. Reference categories are as follows in parentheses for each categorical variable: variant period (Omicron BA.2), age (65+), sex (female), ethnicity (White British), occupation (retired), deprivation (IMD 5 - wealthiest quintile), comorbidities (none), BMI (healthy weight), vaccination status (three doses).

Multiple imputation by chained equations was applied to account for missing covariate data (see Table 1 for missingness) using the *mice* package in R Version 4.0.3 (38), with 5 datasets and 50 iterations per dataset. We conducted a sensitivity analysis using complete cases only to compare with the imputed findings. We also conducted a sensitivity analysis excluding BMI from the adjustment set as it was not required for minimally sufficient adjustment in the final model according to our DAG and had a relatively high proportion of missingness (20%). A further sensitivity analysis was conducted limiting the participants to the sub-cohort who had received serological testing (*n*=4,381, 78%), as testing bias and misclassification of asymptomatic SARS-CoV-2 infections may be less likely to affect this group.

**Table 1.** Features of Study Participants.

| **Characteristic** | **N = 5,630<sup>1</sup>** |
| --- | --- |
| **Age** |  |
| <12 | 151 (2.7%) |
| 12-19 | 141 (2.5%) |
| 20-39 | 290 (5.2%) |
| 40-64 | 2,410 (43%) |
| 65+ | 2,638 (47%) |
| **Sex** |  |
| Female | 3,290 (58%) |
| Male | 2,340 (42%) |
| **Index of Multiple Deprivation Quintile** |  |
| 1 | 379 (6.8%) |
| 2 | 750 (13%) |
| 3 | 1,105 (20%) |
| 4 | 1,497 (27%) |
| 5 | 1,860 (33%) |
| Unknown | 39 |
| **Comorbidities** |  |
| Clinically extremely vulnerable | 729 (13%) |
| Clinically vulnerable | 1,568 (28%) |
| Not clinically vulnerable | 3,333 (59%) |
| **Ethnicity** |  |
| Black | 21 (0.4%) |
| Mixed | 56 (1.0%) |
| Other Asian | 25 (0.4%) |
| Other Ethnicity | 18 (0.3%) |
| South Asian | 65 (1.2%) |
| White British | 5,166 (92%) |
| White Other | 248 (4.4%) |
| Unknown | 31 |
| **Body Mass Index** |  |
| Underweight | 57 (1.3%) |
| Healthy Weight | 1,816 (40%) |
| Overweight | 1,651 (36%) |
| Obesity Class 1 | 651 (14%) |
| Obesity Class 2 | 242 (5.3%) |
| Obesity Class 3 | 108 (2.4%) |
| Unknown | 1,105 |
| **Occupation** |  |
| Higher Risk Occupations | 995 (18%) |
| Lower Risk Occupations | 1,257 (22%) |
| Not in Employment | 241 (4.3%) |
| Retired | 2,444 (43%) |
| Unknown or Other Status | 693 (12%) |

<sup>1</sup>n (%)

## Results

Participant selection is illustrated in Figure 1. Demographic and clinical features of included participants (*n*=5,630) are presented in Table 1. Development of long-term symptoms by infection status and variant period is presented in Supplementary Table S2 and by vaccination status in Supplementary Table S3. Experiencing no infection was the most common infection status across variant periods (*n* range 2428 to 3818). The greatest number of SARS-CoV-2 infections (*n*=959) occurred during the Omicron BA.1 period and the greatest number of other ARIs during the Delta period (*n*=1165).

**Figure 1.**
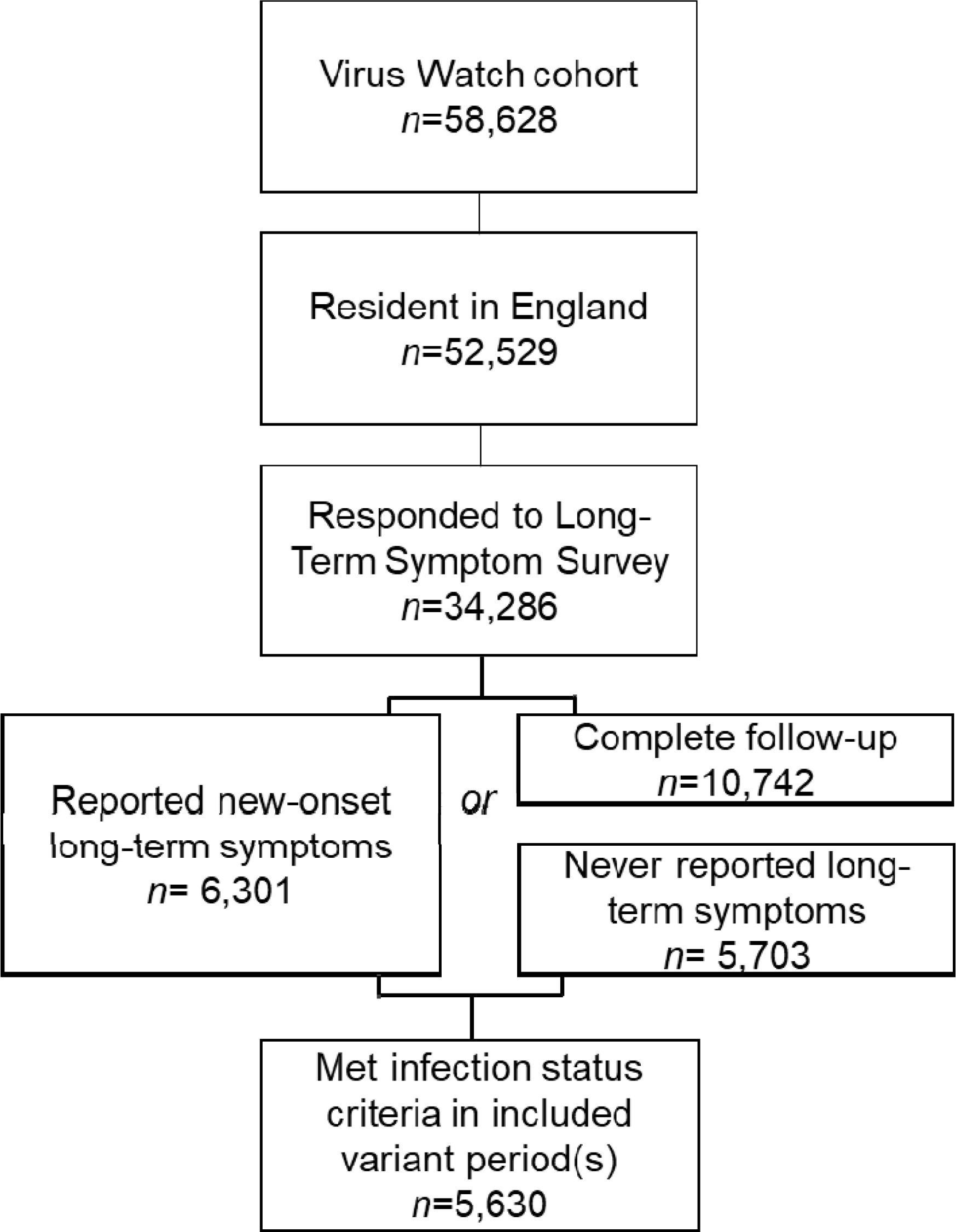
Flow Diagram of Participant Selection.

Predicted probabilities of developing new-onset long-term symptoms meeting the outcome definition are presented by variant dominance period and infection status in Figure 2, with results discussed in light of the study aims below. Odds ratios are presented in Supplementary Table S4.

**Figure 2.**
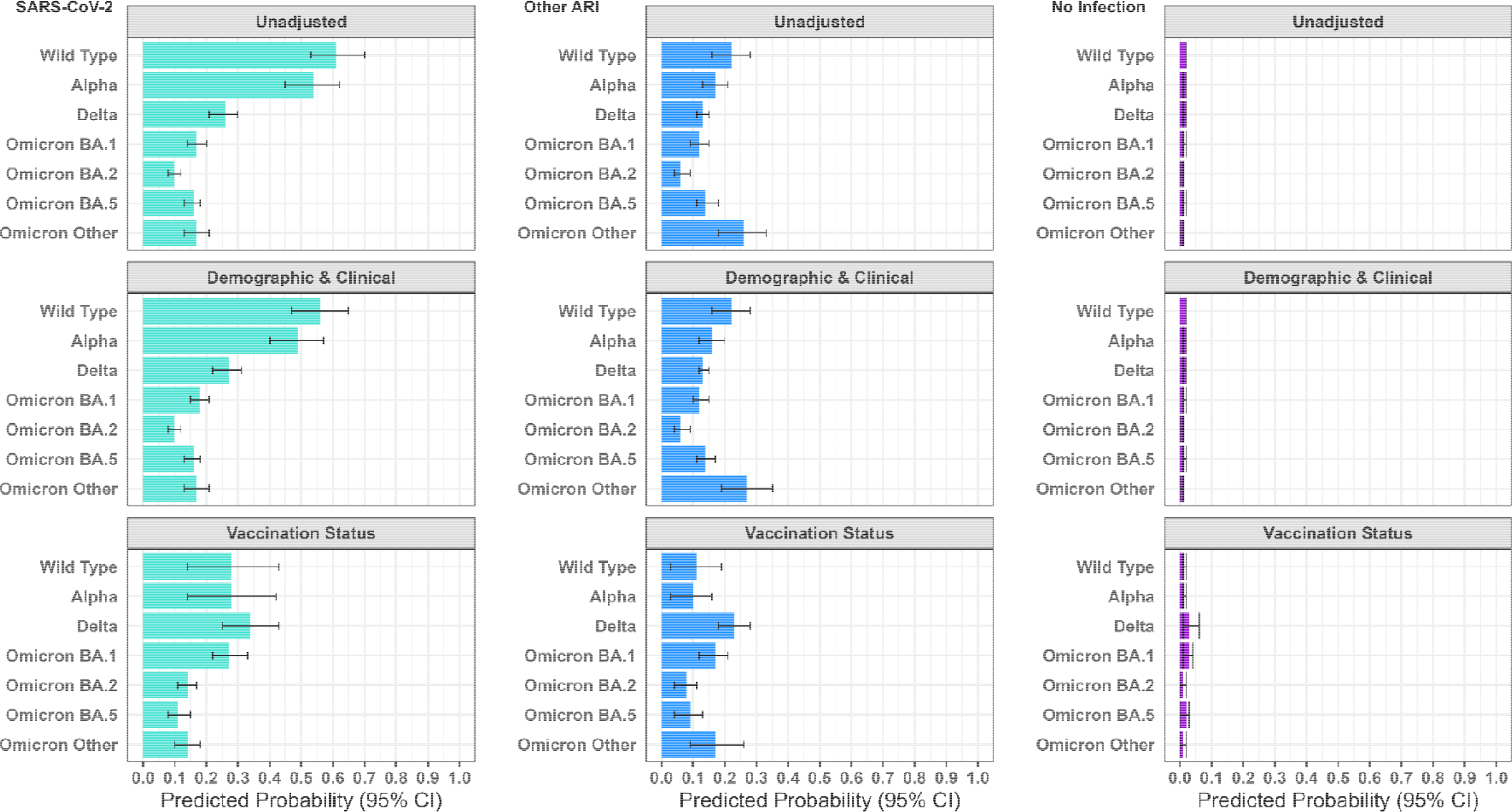
Predicted Probability of New-Onset Long-Term Symptoms by Variant Period and Infection Status.

### SARS-CoV-2 Infections by Variant Period

Predicted probabilities (PP) of developing new-onset long-term symptoms varied substantially by SARS-CoV-2 variant period. In the fully-adjusted model, predicted probabilities for infections during the Wild Type (PP= 0.28, 95% confidence interval (CI) =0.14-0.43), Alpha (PP= 0.28, 95% CI =0.14-0.42), Delta (PP= 0.34, 95% CI =0.25-0.43), and Omicron BA.1 periods (PP= 0.27, 95% CI =0.22-0.33) were elevated compared to all subsequent periods (PP range from 0.11, 95% CI = 0.08-0.15 to 0.14, 95% CI =0.10-0.18).

While differences between variants were identified for SARS-CoV-2 in the fully-adjusted model, these were substantially attenuated compared to earlier models not including vaccination status (see Figure 2). Confidence intervals for the Wild Type and Alpha periods indicated attenuation beyond that expected by chance for the Wild Type (unadjusted PP=0.61, 95% CI =0.53-0.70) and Alpha (unadjusted PP= 0.54, 95% CI =0.45-0.62) periods, which had greater predicted probabilities compared to all other periods in the models excluding vaccination status (unadjusted PP range from 0.10, 95% CI = 0.08-0.12 to 0.26, 95% CI =0.21-0.30).

### Comparison between SARS-CoV-2 and Other Acute Respiratory Infections by Variant Period

In the fully-adjusted model for other ARIs - unlike those for SARS-CoV-2 - no between-period differences were identified beyond those expected due to chance for most variant periods. However, predicted probabilities for the Delta period (0.23, 95% CI 0.18-0.28) - the period with highest estimate - exceeded those for the Omicron BA.2 (0.08, 95% CI 0.04-0.11), Omicron BA.5 (0.09, 95% CI 0.04-0.13), and Alpha (0.10, 95% CI 0.03-0.16).

Compared to models without vaccination status included, confidence intervals indicated a change beyond that expected by chance for the Delta period only following adjustment (unadjusted PP =0.13, 95% CI = 0.11-0.15 versus fully-adjusted PP= 0.23, 95% CI 0.18-0.28); there were also changes in the point estimates for other variant periods, but without evidence of difference beyond that expected due to chance. This may indicate misclassification of SARS-CoV-2 infections as other ARIs in the Delta period.

Estimated predicted probabilities for long-term symptoms following SARS-CoV-2 infection substantially exceeded estimates for other ARIs based on confidence intervals during the Omicron BA.1 period (0.27, 0.22-0.33 for SARS-CoV-2; 0.17, 0.12-0.21 for other ARI). Point estimates for long-term symptoms tended to be higher for SARS-CoV-2 than other ARIs during earlier variant periods, but confidence intervals overlapped. By the Omicron BA.5 and Omicron other periods, estimates were similar for SARS-CoV-2 (PP respectively 0.11, 95% CI = 0.08-0.15 to 0.14, 95% CI =0.10-0.18) and other ARI (PP respectively 0.09, 95% CI = 0.04-0.13 to 0.17, 95% CI =0.09-0.26).

### Comparison between SARS-CoV-2 Infections and No Infection by Variant Period

For the no infection group, the predicted probability of developing long-term symptoms persisting for greater than two months did not differ between variant periods, with predicted probabilities in the fully-adjusted model ranging from 0.01 (95% CI 0.00,0.02) to 0.03 (95% CI 0.01-0.06). There were no within-variant differences beyond those expected by chance across models with different adjustment sets, indicated by overlapping confidence intervals. For all variant periods, the predicted probability of developing long-term symptoms was greater for participants who experienced SARS-CoV-2 infections or other ARIs compared to participants who experienced no infection (Figure 2).

Findings across infection status and variant periods were similar in models based on complete cases (Supplementary Figure S3) and in sensitivity analyses excluding BMI from the adjusted models (Supplementary Figure S4) and including only participants who received serological testing (Supplementary Figure S5).

## Discussion

### Key Findings and Interpretation

Using data from a community prospective cohort in England, this study identified substantial differences in the probability of PCC according to SARS-CoV-2 variant periods. SARS-CoV-2 infection during the Wild Type, Alpha, Delta and Omicron BA.1 periods was associated with greater predicted probabilities (27-34%) of developing long-term symptoms compared to later Omicron sub-variants (11-14%). We also identified differential probability of developing long-term symptoms according to infection status across variant periods. SARS-CoV-2 infection was associated with greater probability of developing long-term symptoms than other ARIs in the Omicron BA.1 period; point estimates were also greater during other early periods but confidence intervals overlapped. Participants who did not experience an infection exhibited stable, low probabilities of developing long-term symptoms (1-3%), which were lower than for participants infected with SARS-CoV-2 or other ARIs.

Findings regarding between-variants difference for SARS-CoV-2 corroborated previous literature indicating that Omicron or Omicron period infections were associated with reduced likelihood of PCC compared to earlier variants (9–20). In the present study, however, the ability to disaggregate Omicron sub-variants found that infection during the Omicron BA.1 period was associated with greater likelihood of PCC compared to subsequent Omicron periods. Overall, findings from this study and the literature indicate that differences in the properties of and consequently response to SARS-CoV-2 variants plausibly influence the likelihood of PCC.

Between-variant differences in the present study persisted after adjustment for sociodemographic and clinical confounding, but these differences were strongly attenuated by adjustment for vaccination status, consistent with previous studies that also accounted for vaccination (19,20). Previous literature regarding the comparative risk associated with the ancestral Wild Type strain had mixed findings, with the current study supporting other adjusted studies which indicated that Wild Type was not more severe than subsequent early VoCs such as Alpha and Delta after accounting for vaccination which became widely available during these later periods (39). The substantial attenuation of between-variant estimates highlights the importance of vaccination as a public health intervention in reducing long-term sequelae as well as short term acute illness in COVID-19, and the importance of accounting for vaccination status in PCC research.

We found evidence of increased odds of developing new-onset long-term symptoms following SARS-CoV-2 infections relative to other ARIs during the Omicron BA.1 and BA.2 periods, with similar differences for other early strains plausible with greater statistical power. The results broadly corroborate previous literature indicating greater likelihood of long-term sequelae following SARS-CoV-2 infection compared to other ARIs during the Wild Type and Alpha periods (24,25). Our study found that for the most recent Omicron sub-variants, however, the likelihood of long-term post-infection sequelae appeared equivalent to other respiratory infections. The trajectory of lower likelihood of PCC in recent Omicron sub-variants is encouraging given their continued dominance. It was not possible to perform genomic or serological testing for non-SARS-CoV-2 infections in the present study.

Therefore, direct comparison with other specific acute respiratory pathogens was not possible and is an area for further investigation. Misclassification of asymptomatic infections with other pathogens into the SARS-CoV-2 group or no infection group is also possible due to the lack of serological testing for other infections, and may have attenuated differences between infection status categories. While adjustment for SARS-CoV-2 vaccination did not alter most estimates for participants who experienced other ARIs or no infection, there was some evidence of misclassification of SARS-CoV-2 cases particularly during the Delta period. This misclassification may have further attenuated differences by infection status. It was also not possible to investigate the impact of co-infections on likelihood of long-term symptoms, which is an increasingly relevant topic given co-circulation of many respiratory pathogens and increased social contact post-pandemic.

Overall, experiencing any acute respiratory infection was associated with substantially greater likelihood of developing new-onset long-term symptoms across all periods compared to not having an infection. These findings correspond with those from an earlier UK study which included only the Alpha variant period (25). While not the central aim of this study, elevated odds of long-term symptoms in all infection groups indicates the relevance of respiratory infections as a risk factor for new-onset long-term symptoms, with some variation across infection type and strain. There may be syndromic differences between symptoms following respiratory infection with different pathogens and those that arise without a preceding infection, but this was beyond the scope of this study and is recommended for future research.

### Strengths and Limitations

This study had several strengths, including the large community cohort and longitudinal design which facilitated collection of long-term symptom data across multiple pandemic periods and facilitated the disaggregation of dominance periods of Omicron sub-variants that were individually designated as VoCs. The combination of testing through national linkage and study records with weekly symptom reporting facilitated direct comparison between SARS-CoV-2 infections, other ARIs, and participants who did not experience infection symptoms. The availability of serological testing for the majority of participants enabled a sensitivity analysis which was less subject to testing and associated detection bias. The WHO consensus definition of PCC was used as the outcome following ARIs and adapted for non-infected participants. Detailed socio-demographic and clinical information collected as part of the Virus Watch study enabled comprehensive adjustment for potential confounding.

However, this study had a number of important limitations. Participants were not fully representative of the English population, and overrepresentation of older, female, and clinically vulnerable participants may have influenced the high overall proportions of participants meeting the definition of PCC. Adjusted predicted probabilities of PCC were, however, in line with previous estimates (1). Participants experiencing long-term symptoms may have been more likely to self-select into completing the questionnaires, likely also influencing overall prevalence though with a less clear effect on differences by infection status and variant period. As self-reporting of long-term symptoms began twelve months after the beginning of the Wild Type period, recall may be less accurate for long-term symptoms beginning during the early phases of the pandemic. Further, 95% of participants had been recruited in 2020 differential reporting bias due to survey fatigue after long-term follow-up may have influenced the lower reporting of long-term symptoms during later Omicron sub-variant periods.

Genomic analysis could not be used to determine the variant and/or pathogen responsible for each infection, and consequently variants may be misclassified and some SARS-CoV-2 infections may be misclassified as other ARIs. This may have attenuated differences by infection status inflating risk of long-term symptoms in other ARIs that were actually attributable to SARS-CoV-2. While a sensitivity analysis limited to participants with serological testing had similar results, asymptomatic SARS-CoV-2 infections may have remained undetected and misclassified into the other ARI and no infection group. The reliance on using symptom reporting to classify other ARIs and no infection meant that follow-up was longer for SARS-CoV-2 during the Wild Type period, as serology and linkage results were available, and likely reduced the other group sizes during the Wild Type period.

While the serological sensitivity analysis aimed to address this, undetected SARS-CoV-2 infections were more likely in the Wild Type period due to more limited community testing. Similarly, SARS-CoV-2 infections may have been under-detected during Omicron sub-variant periods including or occurring after the end of free national testing in April 2022, during which self-testing became a behavioural choice. Although a sub-cohort of Virus Watch study participants were still provided with study-based testing during some parts of this period (see Supplementary Materials) - which may have reduced bias - coverage was not complete and differences by infection status should be cautiously interpreted particularly for the Omicron BA.5 period. However, between-variant differences for SARS-CoV-2 infections - which are limited to those who tested positive - are less likely to be impacted.

Serological assays used in the current study were highly sensitive and specific (please see Supplementary Materials). However, while anti-nucleocapsid antibodies are not the target of current COVID-19 vaccines, serological responses may be less pronounced in vaccinated individuals and understanding of the impact of variants on the natural history of antibody responses is currently limited (40). The impact of these factors on overall detection may be limited however, as the nucleocapsid protein is highly immunogenic and conserved in all SARS-CoV-2 variants and multiple test types were included was likely to reduce related detection bias. Generalisability of the findings to future SARS-CoV-2 variants and to other regions with different vaccination schedules, VoCs that were not present in the UK, and pandemic control measures is uncertain.

### Conclusions

The current study found evidence of elevated likelihood of PCC following infection with SARS-CoV-2 variants up to Omicron BA.1 compared to later Omicron sub-variants. These differences persisted beyond the effects of sociodemographic and clinical confounding, indicating a likely role of between-variant biological differences. However, vaccination substantially attenuated between-variant differences, illustrating its importance as a public health intervention to address elevated risk of long-term sequelae as well as severe acute infection associated with early SARS-CoV-2 variants. Recent Omicron sub-variants demonstrated likelihood of long-term sequelae equivalent to other acute respiratory infections, and having any acute respiratory infection was associated with elevated risk of developing long-term symptoms. Further investigation into the symptomology, burden, and aetiology of long-term post-infection syndromes across SARS-CoV-2 variants and other acute respiratory pathogens is recommended to inform public health and clinical interventions. Notably, aetiological investigations should explicitly consider mechanisms underlying between-variant and between-pathogen differences.

## Funding

Virus Watch was supported by the Medical Research Council [Grant Ref: MC_PC 19070 and MR/V028375/1]. The study also received $15□000 of advertising credit from Facebook to support a pilot social media recruitment campaign on 18 August 2020. The antibody testing was also supported by funding from the Department of Health and Social Care from February 2021 to March 2022. This study was also supported by the Wellcome Trust through a Wellcome Clinical Research Career Development Fellowship to R.W.A. [206602].

From 1 May 2022, Virus Watch received funding from the European Union (END-VoC Project: 101046314). Views and opinions expressed are however those of the author(s) only and do not necessarily reflect those of the European Union or the European Health and Digital Executive Agency (HaDEA). Neither the European Union nor the granting authority can be held responsible for them.

## Competing Interests

AH serves on the UK New and Emerging Respiratory Virus Threats Advisory Group. All other authors declare no competing interests.

## Data Availability

We aim to share aggregate data from this project on our website and via a “Findings so far” section on our website—https://ucl-virus-watch.net/. We also share some individual record level data on the Office of National Statistics Secure Research Service. In sharing the data we will work within the principles set out in the UKRI Guidance on best practice in the management of research data. Access to use of the data whilst research is being conducted will be managed by the Chief Investigators (AH and RA) in accordance with the principles set out in the UKRI guidance on best practice in the management of research data. We will put analysis code on publicly available repositories to enable their reuse.

