## Supplementary Material for "Long-Term Outcomes of SARS-CoV-2 Variants and Other Respiratory Infections: Evidence from the Virus Watch Prospective Cohort in England"

**Long Covid by Variant: Supplementary Material**

**Supplementary Figure S1. Virus Watch Recruitment Dates**

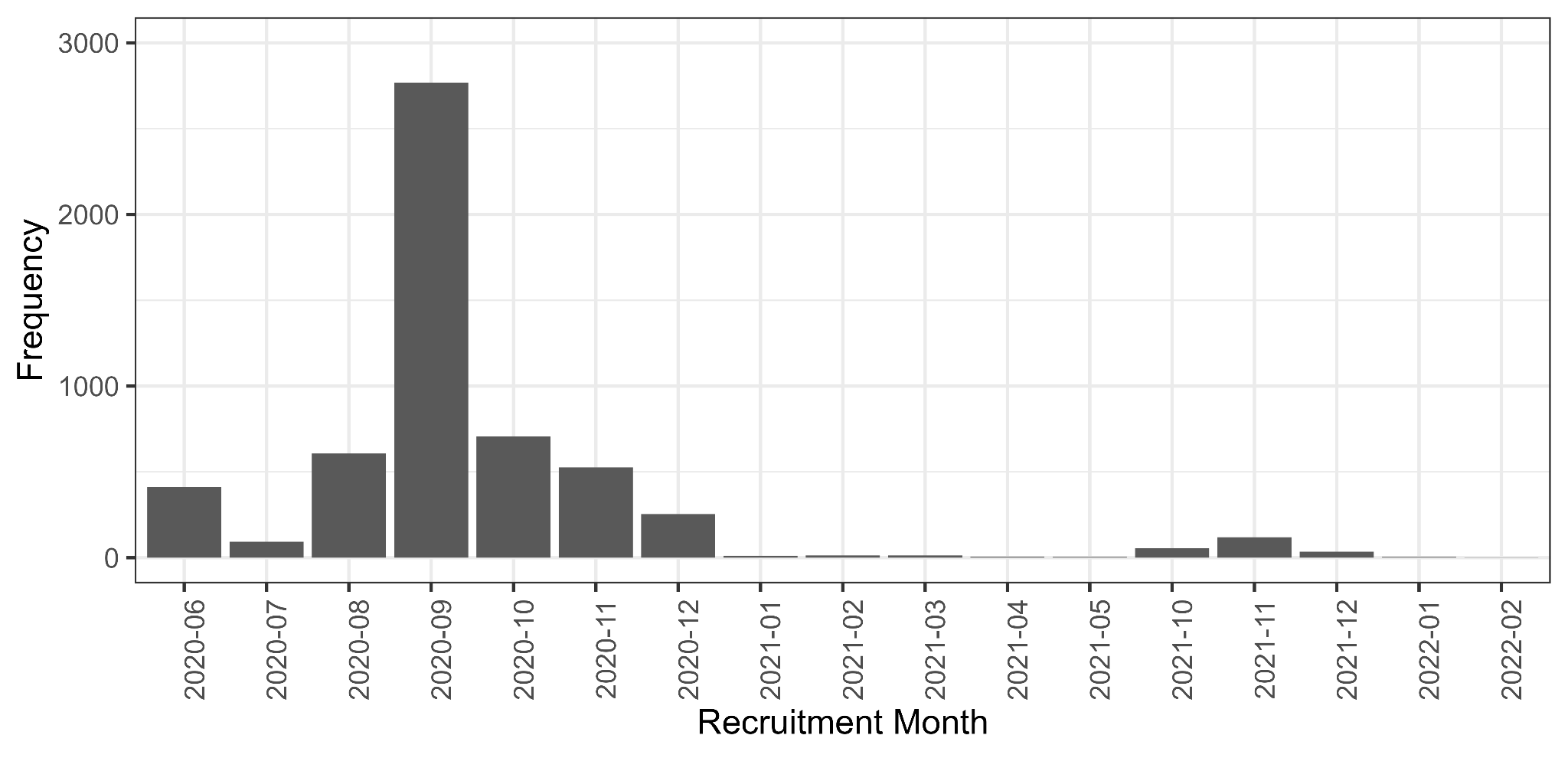

Of participants in the current study, 95% (*n*=5,369) joined the study in the period up to 12-2020. The most common month of recruitment was 09-2020 (*n*=2769, 49%).

**Virus Watch Testing and Study Outcomes**

*Swab-Based Testing (Polymerase Chain Reaction and Lateral Flow Tests)*

Virus Watch testing availability and protocols changed throughout the study. All participants had polymerase chain reaction (PCR) and lateral flow test (LFT) records based on weekly online self-report and linkage to national records across the study period. Between October 2020 and May 2021, a sub-cohort of participants (*n*=1,566 in current study) also posted self-administered PCR nasal swabs for laboratory analysis if symptomatic as part of the Virus Watch study. Between January and July 2023 (after the current study period), a further sub-cohort (*n*=948 in current study) were provided with LFTs to take if symptomatic and report to the study and – if positive – to send a PCR nasal swab for genomic analysis; this analysis is currently underway and genomic results were not available for the current study. However, results around positivity were available and were included in the present data. The study protocol (1) provides further procedural detail regarding swab testing.

*Serological Testing*

Serological testing in Virus Watch was conducted on a sub-cohort of participants using monthly at-home finger-prick serology and/or in-clinic serological testing conducted by healthcare workers. In-clinic serology was performed twice for included participants between September 2020-January 2021 (Autumn round) and April 2021-July 2021 (Spring round) (see study protocol (1) for details). The monthly at-home finger-prick testing was conducted between February 2021 and April 2022. Samples were tested for anti-nucleocapsid and anti-spike antibodies using the Roche Elecsys Anti-SARS-CoV-2 electro-chemiluminescent immunoassays, with estimated sensitivity and specificity of 97% and 99.8% respectively (2). Further detail of testing is provided in the study protocol (1).

As infection date was required to derive the outcome in the current study, only participants with seropositive results that could be attributable to an estimated infection date were included i.e., participants who were initially seronegative and seroconverted during a definable monthly period. Estimated date of infection was defined as the intermediate date between 10 days prior to the last negative, to account for infections that may not have seroconverted, and 14 days prior to the first positive. These were derived based on monthly finger-prick tests. However, serological results from all sources were used to identify participants with evidence of SARS-CoV-2 infection from any source and exclude them where required to prevent misclassification of infection status during each variant period, as described in the Exposure sections of the Methods section.

**Virus Watch Long-Term Symptoms Questionnaire: Symptom list**

**Note**: The symptom list was revised prior to the March 2023 questionnaire to reflect evolving knowledge of post-Covid symptoms and to reduce survey burden by collapsing some items into single items. The ‘Other’ option was available in both questionnaires and the majority of symptoms remained consistent or comparable. Symptom lists are provided here for reference although the outcome in the study comprised any symptom(s) of duration >2 months and (if relevant) within 3 months of infection. Symptom profiles by variant - and consequently symptom harmonisation - are the subject of separate planned analysis beyond the scope of this paper.

**Original Symptom List (February 2021 - March 2022)**

| Fatigue / Feeling unusually tired | Fever |
| --- | --- |
| Headache | Feeling feverish |
| Feeling anxious | Loss of Appetite |
| Feeling depressed/low mood | Trouble Sleeping |
| Runny Nose; Blocked Nose | Lack of concentration |
| Lightheaded or dizzy | Confusion, disorientation |
| Memory loss | Eye redness |
| Dry cough; | Deterioration of eyesight |
| Wet cough | Sore throat |
| Loss of sense of taste | Weakness |
| Altered/distorted sense of taste | Weight loss |
| Loss of sense of smell; | Altered/distorted sense of smell; |
| Reduced sense of smell | Phantom smells in the absence of any odour |
| Hair loss | Chills |
| Rash (all over) | Night Sweats |
| Rash (local) | Sinus pain / congestion |
| Vomiting (being sick) | Sneezing |
| Nausea (feeling sick) | Swollen tonsils |
| Heart Palpitations (Fast or pounding heartbeat) | Wheezing |
| Chest pain (not changed by breathing) | Chest tightness |
| Chest pain when breathing in | Sticky eye |
| Abdominal pain (not inc. menstrual pain) | Eye pain |
| Shortness of breath/difficulty breathing | Ear pain |
| Muscle Aches | Tinnitus (ringing or other noises in one or both ears) |
| Bone or Joint Aches | Change in hearing |
| Other long-term pain | Other symptom(s) |
| Swollen glands |  |

**Symptom List (March 2023)**

| Fatigue | Fever |
| --- | --- |
| Headache | Change in Appetite |
| Feeling anxious | Sleep problems (including difficulty falling asleep, staying asleep, oversleeping or sleep apnoea) |
| Feeling depressed | Brain fog and/or other cognitive problems |
| Congested/runny nose | Dry eyes/ redness of eyes |
| Dizziness | Visual changes; Blurry vision |
| Memory problems | Sore throat or mouth |
| Cough | Post-exertion malaise (feeling unwell or 'crashing' after exercise or physical exertion) |
| Change/loss of taste | Unintentional weight gain |
| Change/loss of smell | Problems with movement, balance, and/or coordination |
| Hair loss | Paraesthesia (pricking, tingling, or creeping feeling on the skin) |
| Skin rash | Swelling of hands and/or feet |
| Unintentional weight loss | Difficulty swallowing (dysphagia) |
| Nausea and/or vomiting | Easy bruising/ bleeding |
| Diarrhoea | Acid reflux/heartburn |
| Tachycardia (fast heartbeat) or palpitations | Gynaecological problems (e.g., change in menstrual cycles or flow) |
| Chest pain | Urinary symptoms (e.g., bladder frequency, urgency or incontinence) |
| Abdominal pain | New allergy (e.g., to food, medications, etc.) |
| Shortness of breath/dyspnoea | Ear pain |
| Muscle aches | Change in hearing |
| Bone and joint pain | Tinnitus |
| Swollen lymph nodes | Other symptom(s), please describe: |

**Supplementary Table S1. Variant Period Dates by English National Region**

| **Region** | **Variant Period** | | | | | |  |
| --- | --- | --- | --- | --- | --- | --- | --- |
|  | **Wild Type** | **Alpha** | **Delta** | **Omicron BA.1** | **Omicron BA.2** | **Omicron BA.5** | **Omicron Other** |
| **East Midlands** | 1 Feb 2020 - 9 Dec 2020 | 4 Jan 2021 - 4 May 2021 | 1 Jun 2021 - 11 Dec 2021 | 20 Dec 2021 - 6 Mar 2022 | 7 Mar 2022 - 12 June 2022 | 4 Jul 2022 - 20 Nov 2022 | < 7 Mar 2023 |
| **East of England** | 1 Feb 2020 - 16 Nov 2020 | 15 Dec 2020 - 27 April 2021 | 25 May 2021 - 11 Dec 2021 | 19 Dec 2021 - 6 Mar 2022 | 7 Mar 2022 - 12 June 2022 | 4 Jul 2022 - 20 Nov 2022 | < 7 Mar 2023 |
| **London** | 1 Feb 2020 - 16 Nov 2020 | 10 Dec 2020 - 13 Apr 2021 | 25 May 2021 - 7 Dec 2021 | 14 Dec 2021 - 6 Mar 2022 | 7 Mar 2022 - 12 June 2022 | 4 Jul 2022 - 20 Nov 2022 | < 7 Mar 2023 |
| **North East** | 1 Feb 2020 - 9 Dec 2020 | 7 Jan 2021 - 11 May 2021 | 1 Jun 2021 - 13 Dec 2021 | 22 Dec 2021 - 6 Mar 2022 | 7 Mar 2022 - 12 June 2022 | 4 Jul 2022 - 20 Nov 2022 | < 7 Mar 2023 |
| **North West** | 1 Feb 2020 - 9 Dec 2020 | 12 Jan 2021 - 20 Apr 2021 | 18 May 2021 - 11 Dec 2021 | 19 Dec 2021 - 6 Mar 2022 | 7 Mar 2022 - 12 June 2022 | 4 Jul 2022 - 20 Nov 2022 | < 7 Mar 2023 |
| **South East** | 1 Feb 2020 - 9 Nov 2020 | 8 Dec 2020 - 27 Apr 2021 | 25 May 2021 - 10 Dec 2021 | 19 Dec 2021 - 6 Mar 2022 | 7 Mar 2022 - 12 June 2022 | 4 Jul 2022 - 20 Nov 2022 | < 7 Mar 2023 |
| **South West** | 1 Feb 2020 - 2 Dec 2020 | 5 Jan 2021 - 13 Apr 2021 | 25 May 2021 - 12 Dec 2021 | 20 Dec 2021 - 6 Mar 2022 | 7 Mar 2022 - 12 June 2022 | 4 Jul 2022 - 20 Nov 2022 | < 7 Mar 2023 |
| **West Midlands** | 1 Feb 2020 - 7 Dec 2020 | 31 Dec 2020 - 11 May 2021 | 25 May 2021 - 12 Dec 2021 | 20 Dec 2021 - 6 Mar 2022 | 7 Mar 2022 - 12 June 2022 | 4 Jul 2022 - 20 Nov 2022 | < 7 Mar 2023 |
| **Yorkshire and the Humber** | 1 Feb 2020 - 16 Dec 2020 | 19 Jan 2021 - 17 May 2021 | 08 Jun 2021 - 12 Dec 2021 | 20 Dec 2021 - 6 Mar 2022 | 7 Mar 2022 - 12 June 2022 | 4 Jul 2022 - 20 Nov 2022 | < 7 Mar 2023 |

**Note:** Missing dates represent time with no variant attributed to >75% of infections; dates for Omicron sub-lineages were only available for England overall; final Omicron Other date represents the closing date of the final survey, with all Omicron dates outside the specified sub-lineages before this date included

**Occupational Categories**

Occupational categories were developed to triangulate occupation, work-related exposure risk, and employment status while preserving statistical power in the current analysis. Occupations had been previously coded in Virus Watch using methodology described elsewhere (1), with categories collapsed to reflect exposure risk based on findings from other Virus Watch studies and previous literature(1)(2,3). The following groups were consequently included in each category:

| **Category** | **Included groups - n (%) of current study population (*N*=5,630)** |
| --- | --- |
| Higher exposure risk occupation | Healthcare  219 (3.9%)  Indoor Trades, Process & Plant  142 (2.5%)  Leisure & Personal Service  99 (1.8%)  Sales & Customer Service  114 (2.0%)  Social Care & Community Protective Services  130 (2.3%)  Teaching, Education & Childcare  250 (4.4%)  Transport & Mobile Machine  41 (0.7%) |
| Lower exposure risk occupation | Administrative & Secretarial  322 (5.7%)  Managers, Directors & Senior Officials  191 (3.4%)  Other Professional & Associate Occupations  687 (12%)  Outdoor Trades  57 (1.0%) |
| Retired | Retired  2,444 (43%) |
| Not in employment | Not in Employment or Homemaker  129 (2.3%)  Permanently sick or disabled  66 (1.2%)  Student  46 (0.8%) |
| Unknown or Other Status | Unknown or Other Status  693 (12%) |

**Supplementary Figure S2. Directed Acyclic Graph for the Impact of Infection Status during each Variant Period on Development of Long-Term Symptoms**

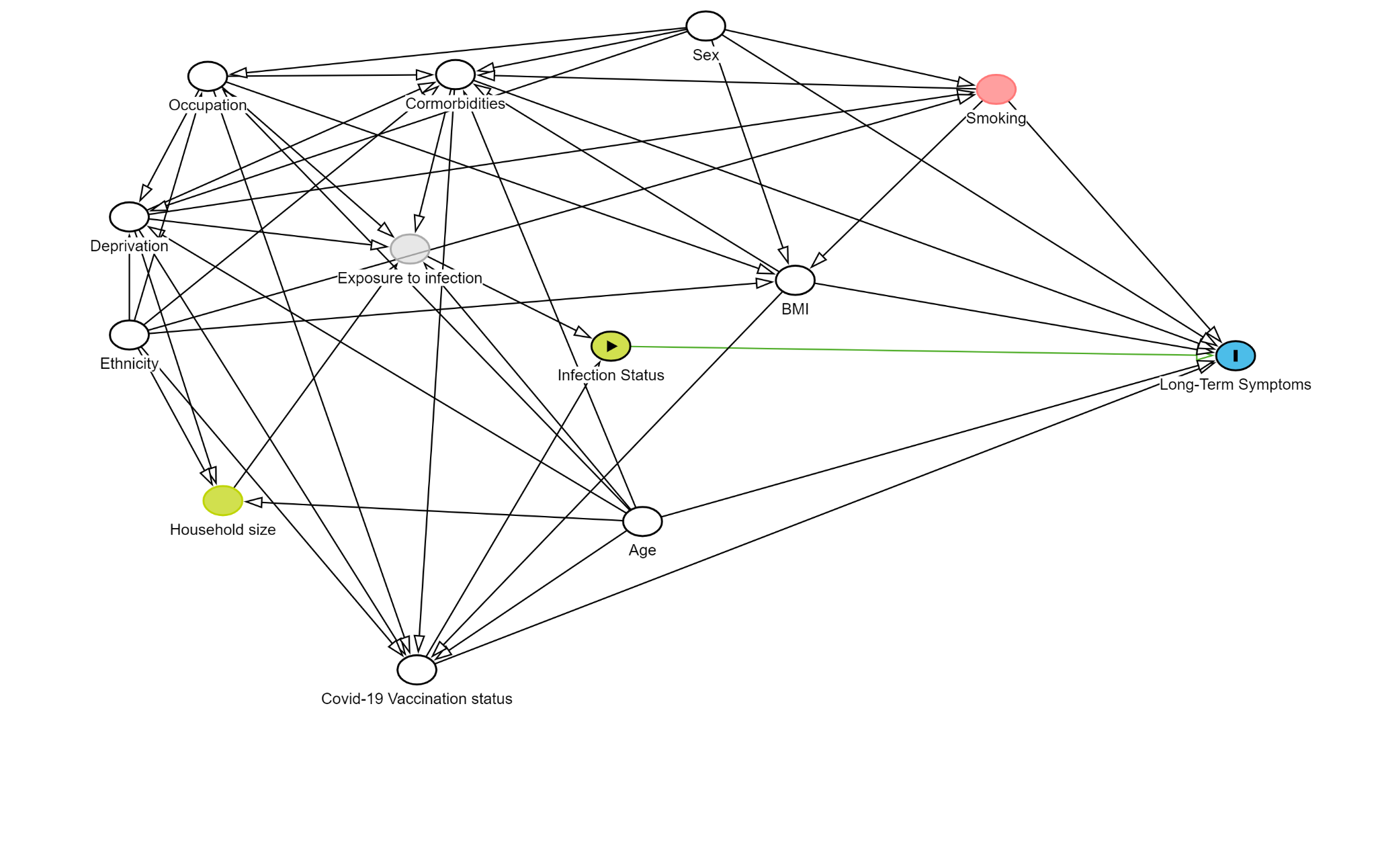

**Supplementary Table S2. Development of New-Onset Long-Term Symptoms by Infection Status and Variant Period**

|  | **SARS-CoV-2** | | | **Other ARI** | | | **No Infection** | | |
| --- | --- | --- | --- | --- | --- | --- | --- | --- | --- |
|  | **Total** | **No Long-Term Symptoms** | **Long-Term Symptoms** | **Total** | **No Long-Term Symptoms** | **Long-Term Symptoms** | **Total** | **No Long-Term Symptoms** | **Long-Term Symptoms** |
| Wild Type | 119 | 46 (39%) | 73 (61%) | 186 | 145 (78%) | 41 (22%) | 3,818 | 3,741 (98%) | 77 (2.0%) |
| Alpha | 127 | 59 (46%) | 68 (54%) | 337 | 279 (83%) | 58 (17%) | 3,597 | 3,530 (98%) | 67 (1.9%) |
| Delta | 434 | 323 (74%) | 111 (26%) | 1,165 | 1,011 (87%) | 154 (13%) | 2,531 | 2,489 (98%) | 42 (1.7%) |
| Omicron BA.1 | 659 | 546 (83%) | 113 (17%) | 522 | 458 (88%) | 64 (12%) | 2,865 | 2,827 (99%) | 38 (1.3%) |
| Omicron BA.2 | 959 | 865 (90%) | 94 (9.8%) | 486 | 455 (94%) | 31 (6.4%) | 2,428 | 2,415 (99%) | 13 (0.5%) |
| Omicron BA.5 | 790 | 666 (84%) | 124 (16%) | 424 | 363 (86%) | 61 (14%) | 2,532 | 2,495 (99%) | 37 (1.5%) |
| Omicron Other | 289 | 240 (83%) | 49 (17%) | 125 | 93 (74%) | 32 (26%) | 2,662 | 2,641 (99%) | 21 (0.8%) |

1 *n* (row %)

**Note:** Participants in the ‘Other ARI’ and ‘No Infection’ groups could be included across multiple variant periods and participants’ infection status and consequently inclusion could change between variant periods; thus, total *n*s for all variant periods vary

**Supplementary Table S3. Development of New-Onset Long-Term Symptoms by Infection Status and Vaccination Status**

|  | | **SARS-CoV-2** | | **Other ARI** | | **No Infection** | |
| --- | --- | --- | --- | --- | --- | --- | --- |
|  | | **No Long-Term Symptoms** | **Long-Term Symptoms** | **No Long-Term Symptoms** | **Long-Term Symptoms** | **No Long-Term Symptoms** | **Long-Term Symptoms** |
| Unvaccinated | | 210 (52%) | 197 (48%) | 477 (80%) | 116 (20%) | 7,591 (97%) | 202 (2.6%) |
| 1 dose | | 57 (80%) | 14 (20%) | 138 (91%) | 13 (8.6%) | 655 (99%) | 6 (0.9%) |
| 2 doses | | 316 (75%) | 105 (25%) | 833 (88%) | 114 (12%) | 2,436 (98%) | 39 (1.6%) |
| 3 doses | | 2,073 (87%) | 322 (13%) | 1,351 (87%) | 207 (13%) | 8,526 (99%) | 85 (1.0%) |
| 4 or more doses | | 225 (81%) | 54 (19%) | 117 (82%) | 25 (18%) | 930 (98%) | 15 (1.6%) |

**Supplementary Table S4. Odds Ratios for New-Onset Long-Term Symptoms by Variant Period and Infection Status (*n*=5,630)**

|  | **Unadjusted – OR (95% CI)** | | | **Demographic & Clinical – OR (95% CI)** | | | **Vaccination Status – OR (95% CI)** | | |
| --- | --- | --- | --- | --- | --- | --- | --- | --- | --- |
|  | **SARS-CoV-2** | **Other ARI** | **No Infection** | **SARS-CoV-2** | **Other ARI** | **No Infection** | **SARS-CoV-2** | **Other ARI** | **No Infection** |
| Wild Type | 14.6 (9.54,22.36) | 4.15 (2.52,6.85) | 3.82 (2.14,6.84) | 13.01 (8.34,20.30) | 4.24 (2.54,7.10) | 3.8 (2.12,6.81) | 2.78 (1.12,6.92) | 1.52 (0.41,5.63) | 1.56 (0.41,5.94) |
| Alpha | 10.61 (7.05,15.96) | 3.05 (1.93,4.82) | 3.53 (1.97,6.30) | 9.51 (6.21,14.56) | 2.89 (1.81,4.61) | 3.49 (1.95,6.24) | 2.73 (1.14,6.54) | 1.32 (0.38,4.57) | 1.45 (0.38,5.48) |
| Delta | 3.16 (2.34,4.28) | 2.24 (1.51,3.31) | 3.13 (1.68,5.85) | 3.44 (2.51,4.73) | 2.31 (1.55,3.44) | 3.04 (1.62,5.68) | 3.79 (2.14,6.72) | 4.22 (2.23,8.01) | 4.16 (1.10,15.71) |
| Omicron BA.1 | 1.9 (1.42,2.55) | 2.05 (1.37,3.08) | 2.5 (1.35,4.63) | 2.03 (1.50,2.74) | 2.08 (1.37,3.14) | 2.43 (1.31,4.53) | 2.61 (1.81,3.76) | 2.66 (1.64,4.32) | 3.26 (1.58,6.76) |
| Omicron BA.2 | REF | REF | REF | REF | REF | REF | REF | REF | REF |
| Omicron BA.5 | 1.71 (1.29,2.28) | 2.47 (1.60,3.81) | 2.75 (1.46,5.19) | 1.73 (1.29,2.31) | 2.46 (1.58,3.82) | 2.74 (1.46,5.17) | 0.77 (0.51,1.16) | 1.15 (0.64,2.05) | 1.94 (0.81,4.63) |
| Omicron Other | 1.88 (1.29,2.73) | 5.05 (2.99,8.52) | 1.48 (0.74,2.95) | 1.88 (1.28,2.75) | 5.69 (3.32,9.73) | 1.45 (0.73,2.90) | 1.01 (0.64,1.59) | 2.84 (1.55,5.19) | 1.06 (0.45,2.50) |

**Note:** REF= reference category

**Supplementary Figure S3.** **Predicted Probability of New-Onset Long-Term Symptoms by Variant Period and Infection Status: Sensitivity analysis using complete cases (*n*=4332)**

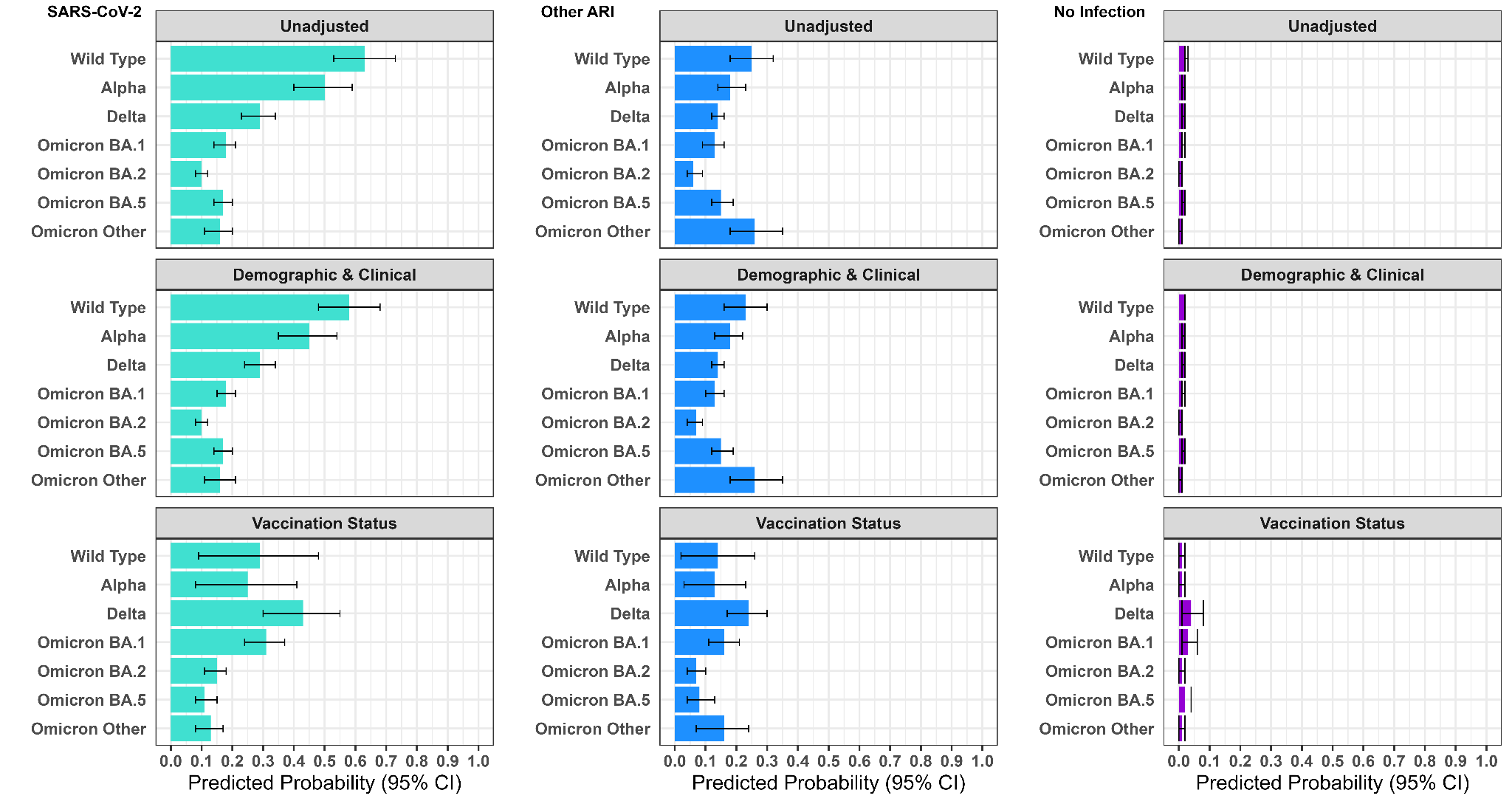

**Supplementary Figure S4.** **Predicted Probability of New-Onset Long-Term Symptoms by Variant Period and Infection Status: Sensitivity analysis excluding body mass index (BMI) from adjusted models (*n*=5,630)**

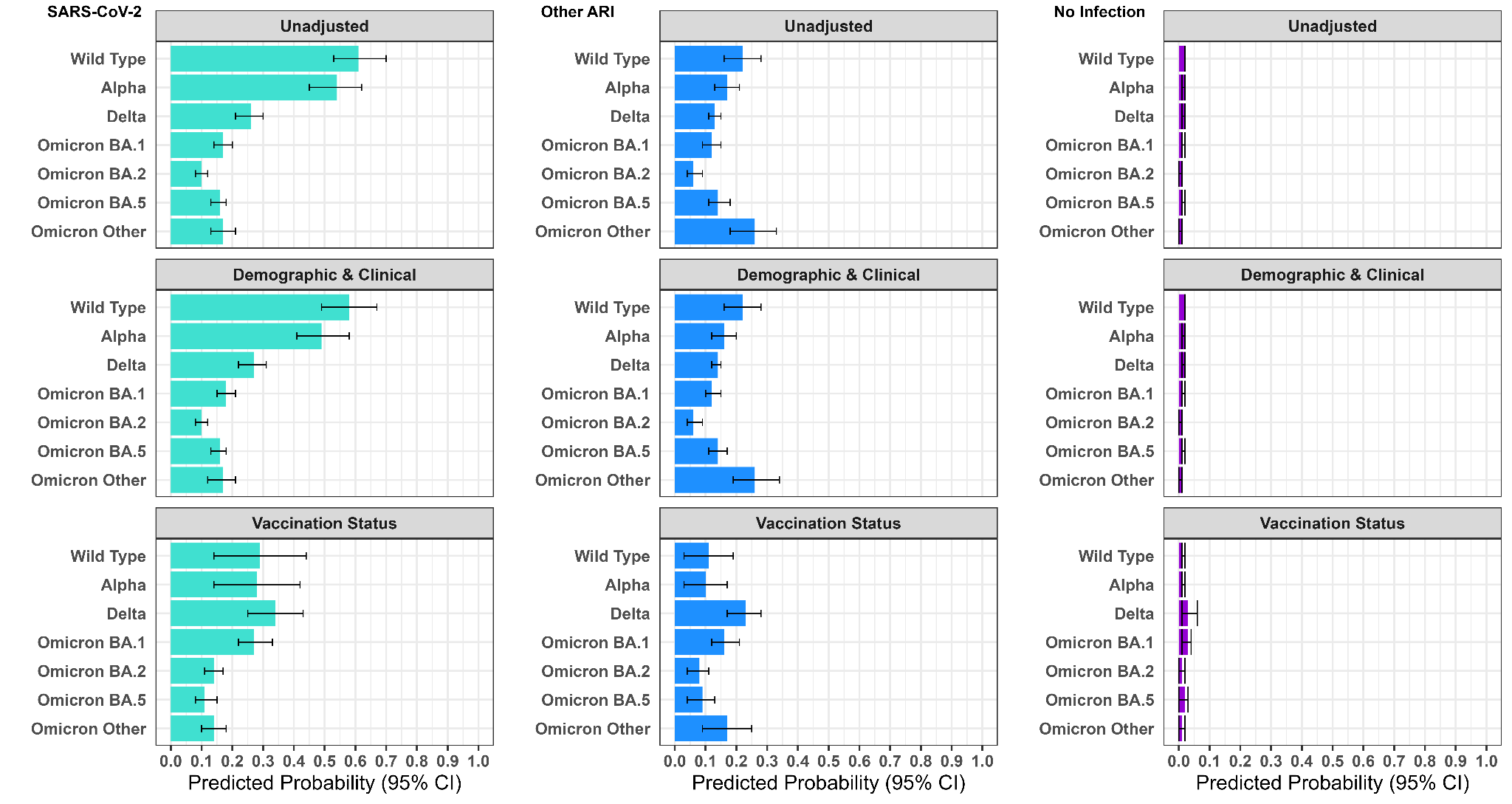

**Supplementary Figure S5.** **Predicted Probability of New-Onset Long-Term Symptoms by Variant Period and Infection Status: Sensitivity analysis including only serological sub-cohort (*n*=4381)**

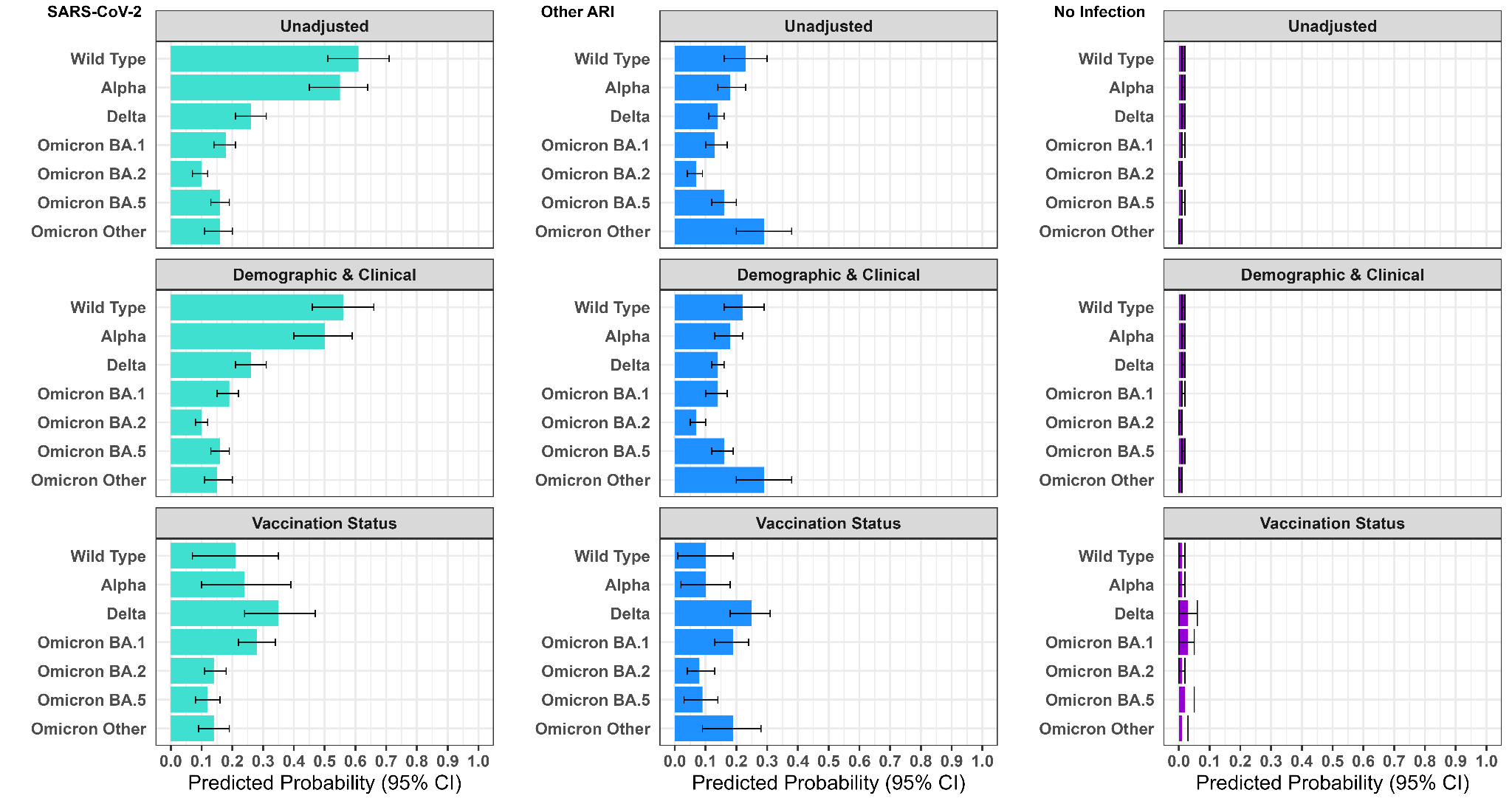
